# COVID-19 and Influenza vaccine co-administration, and missed opportunities to increase COVID-19 vaccination coverage among U.S. adults, 2023-2024

**DOI:** 10.1101/2025.04.14.25325480

**Authors:** Saba A. Qasmieh, Kate Penrose, McKaylee M. Robertson, Rachael Piltch-Loeb, Madhura S. Rane, Avantika Srivastava, Denis Nash

## Abstract

COVID-19 and influenza vaccines are key in protecting against severe clinical outcomes, yet updated COVID-19 vaccination coverage has declined over time and remains much lower than for influenza. Our study aimed to understand the uptake of both COVID-19 and influenza vaccines (including co-administration) and missed opportunities for COVID-19 vaccination during the 2023-2024 respiratory virus season. We surveyed a representative sample of 4022 non-institutionalized U.S. adults and examined COVID-19 and influenza vaccine uptake, and eligibility for and willingness to receive the COVID-19 vaccine. Among all surveyed adults, 19.2% (95% CI 18.0 – 20.5) received both the updated 2023-2024 COVID-19 and influenza vaccines. Co-administration of the influenza vaccine among COVID-19 vaccine recipients was higher (48.5%, 95% CI 45.2 – 51.9) than co-administration of the COVID-19 vaccine among influenza vaccine recipients (22.9%, 95% CI 21.0 – 24.9). Among surveyed adults, 11.2% had received the seasonal influenza vaccine alone and were eligible and willing to receive the COVID-19 vaccine but had not, corresponding to 28.9 million adults who missed an opportunity for COVID-19 vaccination. Given the higher uptake of the influenza vaccine, increasing COVID-19 vaccine co-administration among adults who receive the influenza vaccine could greatly enhance protection against COVID-19 during the respiratory virus season.

## Introduction

COVID-19 and influenza are both associated with substantial morbidity and mortality in the U.S., especially during the late fall and winter^1,2^. The Centers for Disease Control and Prevention (CDC) estimates that there were 21,000 deaths due to influenza and 76,000 COVID-19 deaths in 2022-2023^3,4^. Vaccination against influenza and SARS-CoV-2 viruses remains an effective public health intervention in preventing infection, emergency medical visits, and severe illness leading to hospitalizations and deaths^5–9^. While the risk of SARS-CoV-2 infection in the U.S. remains a year-round threat, the Advisory Committee on Immunization Practices (ACIP) has recommended seasonal influenza and COVID-19 vaccines for the general public that are formulated to protect against co-circulating variants and released in early Fall^10,11^.

While high uptake of vaccination is critical for reducing both the burden on health systems and transmission during respiratory virus season^12,13^, coverage of adult vaccinations remains low. In addition, there are disparities in uptake between influenza and COVID-19 vaccines. In 2022-2023, influenza vaccination coverage was 46.9% among adolescents and adults^14^, compared to 27.1% for the COVID-19 bivalent booster coverage^15^. In December 2023, coverage was 42.2% and 18.3% for the latest influenza and COVID-19 vaccines, respectively^16^.

The ACIP has recommended co-administering the COVID-19 and seasonal influenza vaccines during the same visit among eligible individuals when the timing for each vaccine is appropriate^17^; underscoring the safety and immunogenicity of concomitantly administered influenza and COVID-19 vaccines^18–20^. Co-administration as a vaccination strategy can improve vaccination coverage by reducing missed opportunities to vaccinate^21,22^. While co-administration is considered a widely accepted practice for improving the coverage of childhood vaccinations^23^, uptake of co-administration of adult vaccines, especially during the respiratory virus season, has not been extensively explored or widely promoted as a vaccination coverage improvement strategy for adults in the U.S.^23,24^. Low and disparate coverage between influenza and COVID-19 vaccinations may indicate missed opportunities for vaccination, vaccine hesitancy or both^25^.

Monitoring the uptake of influenza and COVID-19 vaccines is important, however there is limited publicly available population-level data on both vaccines’ co-administration practices. Assessing the landscape of co-administration is an important first step in elucidating potential strategies to improve vaccination coverages for both respiratory pathogens. The objective of this study was to assess the COVID-19 and seasonal influenza vaccination coverage and administration practices among U.S. adults during the 2023-2024 respiratory virus season. We further examined the uptake of the COVID-19 vaccine among influenza vaccine recipients, a potential missed opportunity for COVID-19 vaccination, and assessed the potential impact of co-administration as an implementation strategy for improving COVID-19 vaccination coverage.

## Methods

### Study Population

Between January 19^th^ and 28^th^ 2024, Ipsos conducted a cross-sectional population-representative survey (in English and Spanish) among U.S. adults 18 years and older. The sample was drawn using an equal probability of selection method from the national opt-in Ipsos KnowledgePanel of 55,000 non-institutionalized US adults^26^. Survey weights were created using a raking approach to adjust for differential nonresponse and to ensure representativeness of the U.S. adult population.

### Study Definitions

#### Vaccination coverage

Coverage for the updated 2023-2024 COVID-19 vaccine (hereafter COVID-19 vaccine) was defined based on the proportion of adults who reported receiving a COVID-19 vaccine since September 11, 2023, when the monovalent vaccine targeting the XBB.1.5 variant was authorized^27^. Influenza vaccination coverage was defined based on the proportion of adults who reported receiving an influenza vaccine since July 1, 2023, when influenza vaccines were first available for the 2023-2024 respiratory season.

#### Vaccine co-administration

Co-administration was defined based on adults who reported receiving the seasonal influenza vaccine on the same day as their last COVID-19 vaccination. Adults who reported receiving both the influenza and the COVID-19 vaccines but not on the same day were classified as having received both vaccines separately.

#### Missed opportunity for COVID-19 vaccination

The World Health Organization and the CDC define missed opportunity for vaccination (MoV) as times when individuals who are eligible for a vaccine have contact with healthcare services but do not receive a vaccine^28,29^. Given the higher coverage of seasonal influenza vaccination compared to the updated COVID-19 vaccine, we define adults who had a missed opportunity for COVID-19 vaccination as those who had contact with the healthcare system to receive the influenza vaccine only. Among those who received the influenza vaccine, we further classified adults who were both eligible for and willing to receive the COVID-19 vaccine. Eligibility to receive the COVID-19 vaccine was limited to those who reported their most recent SARS-CoV-2 infection before September 2023 (i.e., before the 2023-2024 COVID-19 dose was approved). Willingness to receive the COVID-19 vaccine was defined based on adults who reported they were very or somewhat willing to get the annual COVID-19 shot if it was recommended by their healthcare provider or doctor.

#### Socio-demographic characteristics

The following information was collected on survey respondents: age, gender, race/ethnicity, education, household income, employment status, geographic region, health insurance, access to healthcare provider, and any pre-existing chronic conditions which was defined as having one of the following: diabetes, chronic obstructive pulmonary disease (COPD) or lung disease, cystic fibrosis, tuberculosis, asthma, liver disease, heart disease, high blood pressure, cerebrovascular disease, chronic kidney disease, depression, anxiety, another mental health condition, dementia, or an immunocompromising condition (i.e., a cancer, recent organ transplant, HIV, or taking immunosuppressive medications). Reported barriers to care was defined as those who reported one of the following: did not have health insurance, did not have a personal doctor or healthcare provider, or they needed to see a doctor in the past 12 months but did not because of cost. The study protocol was approved by the Institutional Review Board at the City University of New York (CUNY IRB 2022–0407).

### Statistical Analysis

Vaccination coverage was calculated as a weighted proportion of all U.S. adults who received 1) COVID-19 vaccine overall (with or without influenza vaccine), 2) influenza vaccine overall (with or without COVID-19 vaccine), 3) only the COVID-19 vaccine (without influenza vaccine), 4) only the influenza vaccine (without COVID-19 vaccine), and 5) both COVID-19 and influenza vaccines.

We estimated the weighted number of U.S. adults using 2019-023 American Community Survey 5-year estimates using an inference population of 258,742,302 adults^30^. We examined the proportion who received a co-administered COVID-19 vaccine among influenza vaccine recipients, co-administered influenza vaccine among COVID-19 vaccine recipients, and co-administration of both vaccines among recipients of both COVID-19 and influenza vaccines (as opposed to receipt of vaccines at separate visits). We generated 95% confidence intervals (CI) for each coverage and for co-administration.

Frequencies and weighted proportions were generated for the following vaccination groups: not yet vaccinated for either respiratory pathogen, received only the COVID-19 vaccine, received only the seasonal influenza vaccine, and received both vaccines (at separate and same visit). Estimates were stratified by select socio-demographic characteristics. We performed a Pearson’s Chi-square test of independence to examine vaccination group differences. Co-administration of COVID-19 vaccine among influenza vaccine recipients was examined by key population only, namely age, race/ethnicity, pre-existing chronic conditions and barrier to healthcare status. We generated the weighted proportion of COVID-19 vaccine eligible and willing adults among influenza-only vaccine recipients. We calculated estimated change in COVID-19 vaccination coverage by summing the number adults who already received the COVID-19 vaccine and those could have received the COVID-19 vaccine but did not due to MoV among all adults, and then stratified further by age, race/ethnicity, pre-existing conditions, and with barriers to care. Data analyses were conducted using SAS, version 9.4 and R Studio.

## Results

Among 4022 U.S. adults, coverage for the COVID-19 vaccine was 21.1% (95% CI 19.9 – 22.5) and 44.9% (95% CI 43.4 – 46.5) for the influenza vaccine. Among all adults, 1.9% (95% CI 1.5 – 2.4) received the COVID-19 and not the influenza vaccine and 25.6% (95% CI 24.2 – 27.0) received the influenza vaccine and not the COVID-19 vaccine. Coverage of both COVID-19 and influenza vaccinations was 19.2% (95% CI 18.0 – 20.5). Among adults vaccinated against both COVID-19 and influenza, 53.4% (95% CI 50.0 – 56.9) received both vaccines during the same visit. Among influenza vaccine recipients, 22.9% (95% CI 21.0 – 24.9) received the COVID-19 vaccine during the same visit, and among COVID-19 vaccine recipients, 48.5% (95% CI 45.2 – 51.9) received the influenza vaccine during the same visit (Figure 1).

**Figure 1:**
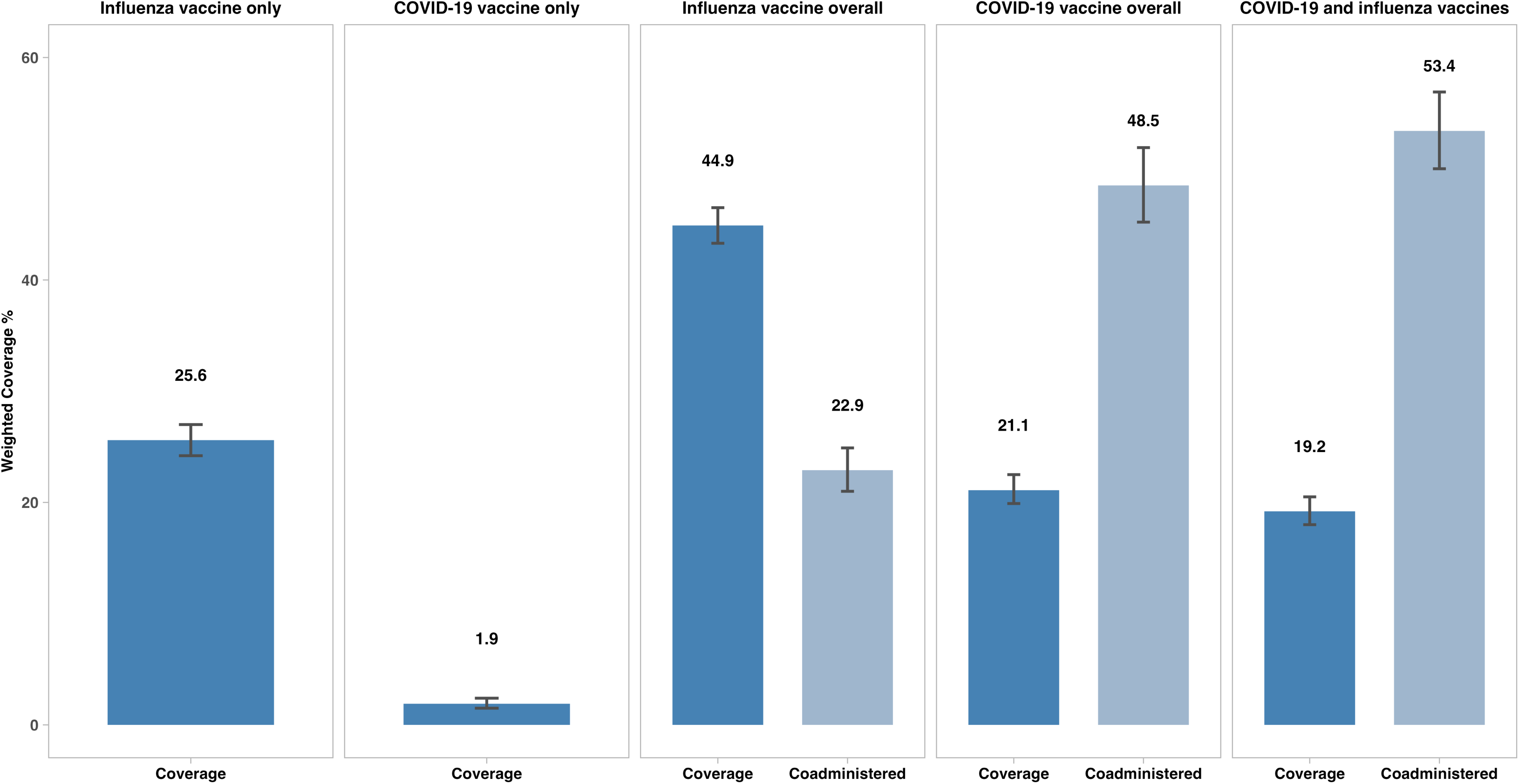
Coverage and co-administration of the seasonal influenza and the updated 2023-2024 COVID-19 vaccines, 2023-2024 respiratory virus season, United States.

Adults unvaccinated against both pathogens were commonly adults between 18-44 years (65.0%, p-value <0.0001), Hispanic adults (62.8%, p-value <0.0001), those without insurance (88.1%, p-value <0.0001), those with a pre-existing condition (60.9%, p-value <0.0001), and those with barriers to care (72.9%, p-value <0.0001) (Table 1). Vaccination against both COVID-19 and influenza was higher among adults ≥65 years (33.1%, p-value <0.0001) than adults 18-44 years (12.2%) and 45-64 years (19.3%). Vaccination against both COVID-19 and influenza was also more common among adults with a Bachelor’s degree or higher (31.6%) compared to those with high school level education or below (9.7%, p-value <0.0001). More adults with health insurance were vaccinated with both vaccines (21.1%) compared to adults without insurance (2.2%, p-value <0.0001). A higher proportion of adults with a medical provider were vaccinated with both COVID-19 and influenza vaccines (22.8%) compared to those without (9.2%, p-value <0.0001).

**Table 1:** Socio-demographic differences across uptake of the 2023-2024 COVID-19 and influenza vaccines, 2023 - 2024 respiratory virus season, United States.

| Characteristics | Total | Unvaccinated<sup>a</sup> | Received COVID-19 vaccine only | Received Influenza vaccine only | Received both COVID-19 and influenza vaccines | P-value<sup>b</sup> |
| --- | --- | --- | --- | --- | --- | --- |
|  | N (%) | N (%) | N (%) | N (%) | N (%) |  |
|  | 4022 (100) | 2139 (53.2) | 78 (1.9) | 1030 (25.6) | 775 (19.3) |  |
| **Age** |  |  |  |  |  |  |
| 18–44 | 1853 (46.1) | 1204 (65.0) | 27 (1.4) | 392 (21.2) | 230 (12.2) | <0.0001 |
| 45 - 64 | 1254 (32.2) | 668 (53.2) | 27 (2.2) | 318 (25.3) | 242 (19.3) |  |
| ≥65 years | 914 (22.7) | 267 (29.2) | 24 (2.6) | 321 (35.1) | 302 (33.1) |  |
| **Gender** |  |  |  |  |  |  |
| Man | 1970 (49.0) | 1067 (54.2) | 38 (1.9) | 483 (24.5) | 383 (19.4) | 0.433 |
| Woman | 2051 (51.0) | 1071 (52.2) | 40 (1.9) | 548 (26.7) | 392 (19.1) |  |
| **Race/ethnicity** |  |  |  |  |  |  |
| Black NH | 486 (12.1) | 261 (53.8) | 12 (2.4) | 138 (28.5) | 74 (15.2) | <0.0001 |
| Hispanic | 704 (17.5) | 442 (62.8) | 18 (2.6) | 148 (21.1) | 95 (13.5) |  |
| White NH | 2464 (61.3) | 1257 (51.0) | 42 (1.7) | 645 (26.2) | 521 (21.1) |  |
| Other NH | 368 (9.1) | 178 (48.4) | 5 (1.5) | 99 (26.9) | 85 (23.2) |  |
| **Education** |  |  |  |  |  |  |
| HS graduate or less | 1536 (38.2) | 992 (64.5) | 27 (1.7) | 369 (24.0) | 149 (9.7) | <0.0001 |
| Some college | 1062 (26.2) | 594 (55.9) | 23 (2.2) | 263 (24.7) | 182 (17.1) |  |
| Bachelor's degree or above | 1423 (35.4) | 553 (38.8) | 28 (1.9) | 399 (28.1) | 444 (31.6) |  |
| **Household income** |  |  |  |  |  |  |
| Below \$50,000 | 1064 (26.4) | 660 (62.0) | 20 (1.9) | 275 (25.8) | 109 (10.3) | <0.0001 |
| \$50,000 - \$99,999 | 1142 (28.4) | 645 (56.5) | 19 (1.7) | 279 (24.5) | 198 (17.4) | |
| \$100,000 or above | 1816 (45.2) | 834 (45.9) | 38 (2.1) | 477 (26.3) | 467 (25.7) | |
| **Currently employed** |  |  |  |  |  |  |
| Yes, full-time | 1905 (47.4) | 1079 (56.7) | 37 (2.0) | 455 (23.9) | 333 (17.5) | <0.0001 |
| Yes, part-time | 564 (14.0) | 310 (55.0) | 18 (3.2) | 128 (22.7) | 108 (19.1) |  |
| No | 1553 (38.6) | 750 (48.3) | 22 (1.4) | 447 (28.8) | 334 (21.5) |  |
| **Geographic region** |  |  |  |  |  |  |
| Northeast | 698 (17.4) | 328 (47.0) | 13 (1.8) | 211 (30.2) | 146 (21.0) | <0.0001 |
| Midwest | 823 (20.5) | 447 (54.4) | 11 (1.4) | 195 (23.7) | 169 (20.6) |  |
| South | 1551 (38.6) | 885 (57.1) | 22 (1.4) | 399 (25.7) | 245 (15.8) |  |
| West | 950 (23.6) | 478 (50.3) | 32 (3.3) | 226 (23.8) | 214 (22.5) |  |
| **At least one pre-existing condition<sup>c</sup>** |  |  |  |  |  |  |
| Yes | 2212 (55.0) | 1024 (46.3) | 30 (1.3) | 657 (29.7) | 501 (22.6) | <0.0001 |
| No | 1730 (43.0) | 1053 (60.9) | 47 (2.7) | 367 (21.2) | 263 (15.2) |  |
| **Barriers to healthcare<sup>d</sup>** |  |  |  |  |  |  |
| Yes | 1274 (31.7) | 929 (72.9) | 22 (1.7) | 205 (16.1) | 118 (9.3) | <0.0001 |
| No | 2748 (68.3) | 1209 (44.0) | 56 (2.0) | 826 (30.1) | 657 (23.9) |  |
| **Insurance** |  |  |  |  |  |  |
| Yes, has insurance | 3627 (90.2) | 1791 (49.4) | 73 (2.0) | 997 (27.5) | 766 (21.1) | <0.0001 |
| No, not insured | 394 (9.8) | 348 (88.1) | 5 (1.2) | 34 (8.5) | 9 (2.2) |  |
| **Was there a time you could not see a doctor in the last 12 months** |  |  |  |  |  |  |
| Yes | 319 (7.9) | 233 (73.0) | 2 (0.8) | 62 (19.6) | 21 (6.7) | <0.0001 |
| No | 3703 (92.1) | 1906 (51.5) | 75 (2.0) | 969 (26.2) | 754 (20.4) |  |
| **Has a medical provider** |  |  |  |  |  |  |
| Yes | 2992 (74.4) | 1363 (45.6) | 58 (1.9) | 890 (26.8) | 680 (22.8) | <0.0001 |
| No | 1030 (25.6) | 775 (75.2) | 20 (2.0) | 141 (13.7) | 94 (9.2) |  |

Compared to adults without any barriers to healthcare services (23.9%), fewer adults with barriers to healthcare access were vaccinated with influenza and COVID-19 vaccines (9.3%, p-value <0.0001) (Table 1).

Among influenza vaccine recipients overall, non-Hispanic Black adults more commonly reported receiving only the influenza vaccine compared to adults of other racial/ethnic groups (Figure 2). More adults ages 18-34 and 45-54 years old (64.0%, 58.4%, respectively) received only the influenza vaccine and not the COVID-19 vaccine than adults ≥65 years (51.4%, p-value <0.0001). More adults ≥65 years who received the influenza vaccine received the COVID-19 vaccine on different days (27.9%, p-value <0.0001) than on the same day (20.7%). The proportion of adults ≥65 years who received both vaccines, but on separate days, was higher than any other age group. Among influenza vaccine recipients, receipt of the COVID-19 vaccine on separate days was highest among adults without barriers to care (21.6%) compared to those with barriers (12.7%, p-value <0.0001) (Figure 2).

**Figure 2:**
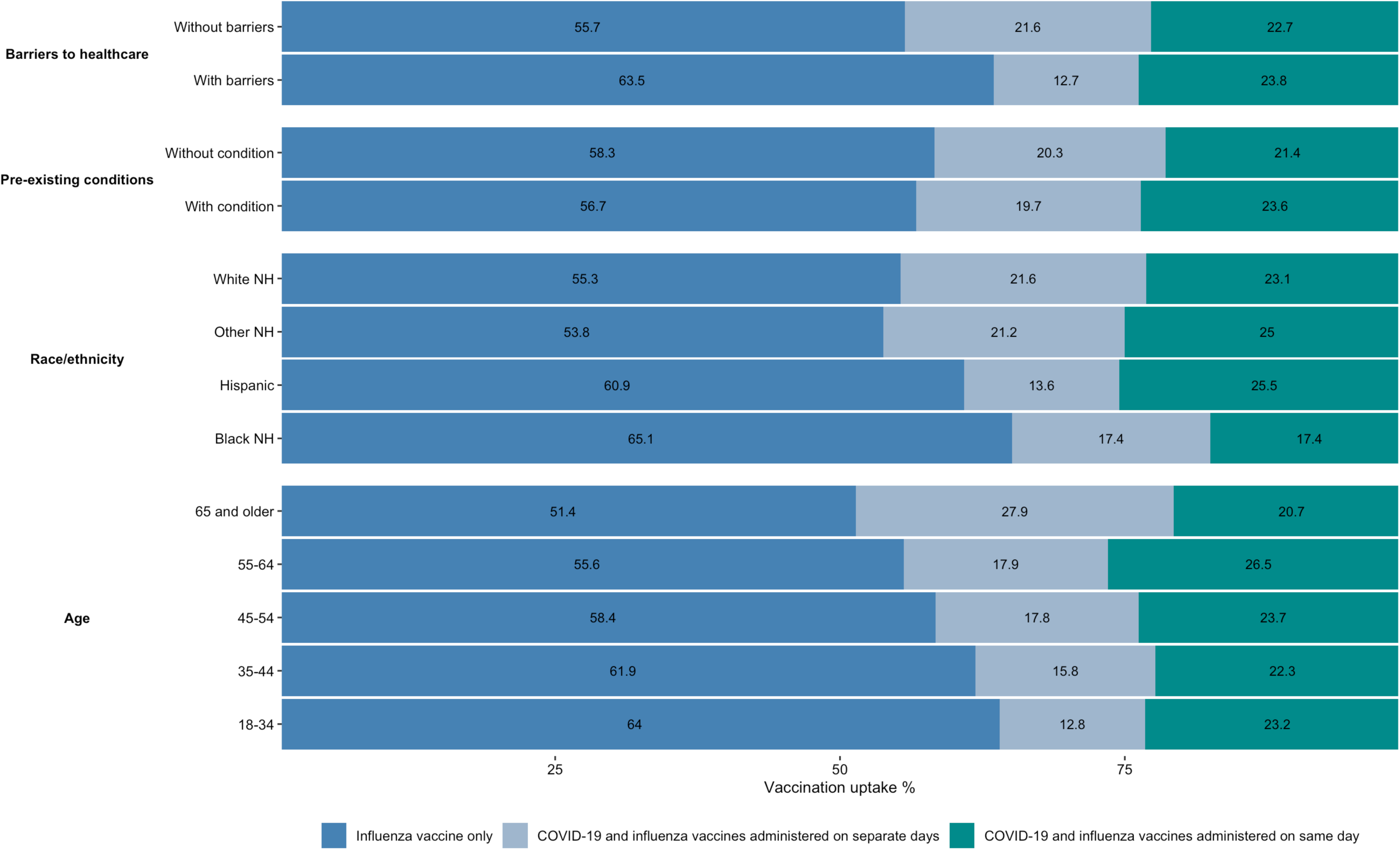
COVID-19 vaccination uptake among influenza vaccination recipients across age, race/ethnicity, pre-existing chronic conditions and barriers to care, 2023-2024 respiratory virus season, United States.

Among the 1031 (25.6%) surveyed adults who only received the influenza vaccine, 898 (87.1%) did not report having a SARS-CoV-2 infection during the study period and were eligible to receive the COVID-19 vaccine, and half of those eligible, 450 (50.1%), reported willingness to receive the COVID-19 vaccine. Among COVID-19 vaccine-eligible influenza-only vaccine recipients, willingness was highest among young adults 18-34 years (53.1%) and Hispanic adults (57.4%). Vaccinating influenza-only vaccine recipients who missed an opportunity to receive the COVID-19 vaccine was estimated to increase overall COVID-19 vaccination coverage from 20.8% to 32.0%, an 11.2%-point coverage increase among all adults. Vaccination of adults ≥ 65 years who were both eligible and willing to receive the COVID-19 vaccine would increase COVID-19 vaccination coverage to 51.1% in this age group, a 15.8%-point coverage increase. Further, COVID-19 vaccination of adults who missed an opportunity to vaccinate against COVID-19 would lead to major increases in coverage among non-Hispanic Black adults (31.3%, a 14.0%-point coverage increase) and adults with a pre-existing condition (36.5%, a 13.0%-point coverage increase) (Table 2)

**Table 2:**
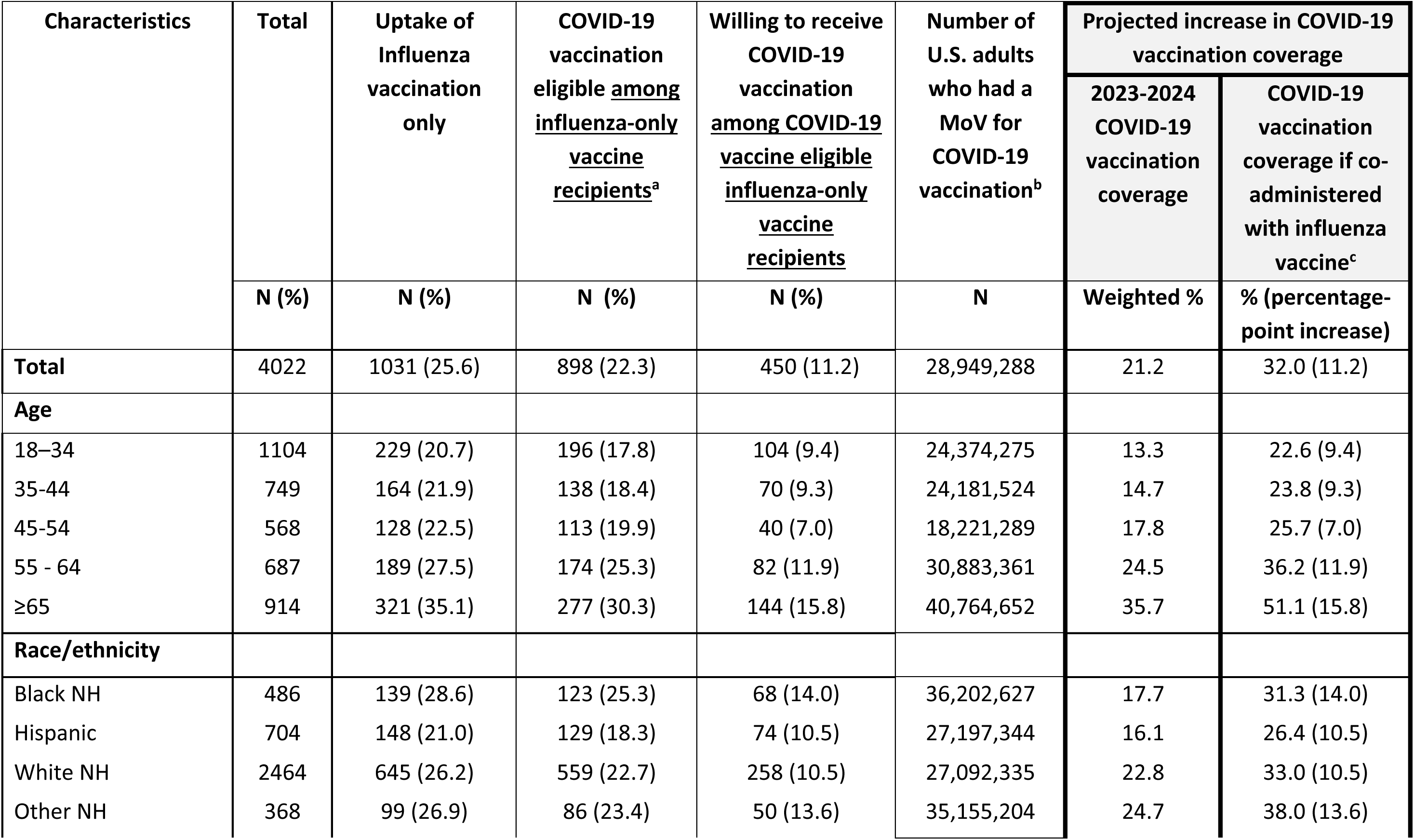
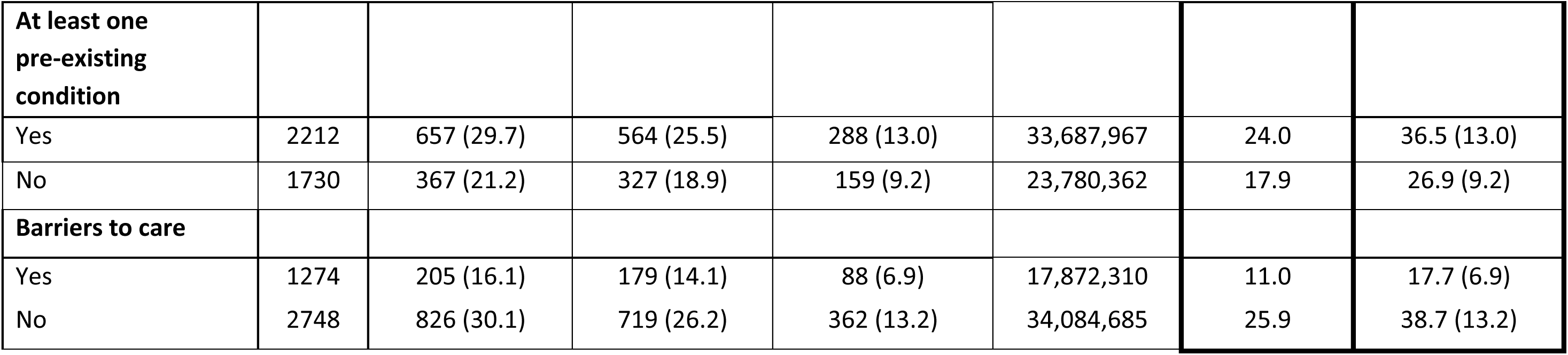
Missed opportunity to vaccinate (MoV) against COVID-19 and estimated change in COVID-19 vaccination coverage among COVID-19 eligible and willing influenza-only vaccine recipients, 2023-2024 respiratory virus season, United States.

## Discussion

We observed low COVID-19 and seasonal influenza vaccination coverages during the 2023-2024 respiratory virus season in this population-representative sample. Influenza vaccination coverage, while higher than COVID-19 vaccination coverage, was well below the Healthy People 2030 target of 70% among all adults^31,32^. Our coverage estimates for both influenza and COVID-19 vaccination aligned with national coverage estimates published by the CDC during the same time period^33^, which did not substantially change by the end of May 2024. We observed disparities in vaccination coverage across socio-demographic characteristics and higher coverage of both vaccinations among adults with higher socioeconomic characteristics and better access to care. Additionally, a substantial proportion of adults who only received the influenza vaccine had a missed opportunity for COVID-19 vaccination.

Higher coverage of influenza vaccination relative to COVID-19 could be explained by wider acceptance of the influenza vaccine, which has been licensed and available in the United States since 1945^34^, and a public more accustomed to receiving an annually updated vaccine to match circulating influenza subtypes^21^. As such, and given that it has been available for longer periods of time than the COVID-19 vaccine, the seasonal influenza vaccine is more commonly and routinely recommended for high-risk groups such as older adults than the COVID-19 vaccine. Studies have also shown higher vaccine hesitancy towards COVID-19 vaccines relative to other vaccines^35^. In our study, approximately half of adults who received the influenza vaccine and were eligible to receive the COVID-19 vaccine expressed willingness to receive the COVID-19 vaccine. Unwillingness among the other half of willing adults suggests hesitancy towards COVID-19 vaccine recommendations even among eligible adults, and the need for targeted and culturally sensitive communication and messaging around the benefits of COVID-19 vaccines that are tailored to public perceptions^25^.

About half of surveyed adults who were vaccinated against COVID-19 also received the influenza vaccine during the same visit. This estimate was similar to a study based on registrants using CDC’s vaccine safety monitoring system, V-safe, in October 2023^23^. We observed high coverage of both influenza and COVID-19 vaccinations (on same or separate visits) among adults ≥65 years, a key age group associated with increased morbidity and mortality due to both pathogens. Even though coverage for both COVID-19 and influenza vaccination was highest among adults ≥65 years, adults in this age group more commonly received both vaccines on separate days than during the same visit. Non-concomitantly administered vaccines among adults ≥65 years may point to accessibility to comprehensive healthcare coverage (e.g. Medicare) and thus improved coverage for and access to preventative services. As such, more care encounters and fewer out of pocket expenses may lead to fewer MoV. We observed similar trends among adults without barriers to care who commonly reported receiving both COVID-19 and influenza vaccines separately. A higher propensity of adults ≥65 years receiving both vaccines at separate visits may also be attributed to having concerns of adverse effects due to prior co-administration of other adults vaccines^36^, or due to vaccine timing (e.g., may have received influenza vaccine before the start of influenza and opted to wait until COVID-19 vaccines were available to get them at separate visits).

The seasonal influenza vaccine is the most common vaccine to be potentially co-administered with the COVID-19 vaccine due to the vaccine being commonly recommended, has broad eligibility, and/or due to influenza co-circulating at the same time as SARS-CoV-2 during the respiratory season. It can, therefore, be viewed as an entry point to improve COVID-19 vaccination and other adult vaccines if the timing for those vaccines is right. In our study, COVID-19 vaccine recipients were more likely to also receive the influenza vaccine than influenza vaccine recipients receiving the COVID-19 vaccine. By identifying those adults as a key target group, co-administration of the COVID-19 vaccine during the influenza vaccine visit could have resulted in an increase in COVID-19 coverage, particularly among vulnerable groups such as older adults and among racial/ethnic minority groups. Co-administration may also be an effective strategy targeting adults not vaccinated against either vaccine and who have reported having barriers to healthcare.

Studies have shown that acceptability of concomitant administration of the COVID-19 and influenza vaccines has increased since 2021 when the ACIP first recommended COVID-19 vaccine to be administered with other vaccines regardless of timing^23,37^. In our study, more than half of adults who received both vaccines during the 2023-2024 season received both during the same visit. Given that peak season to vaccinate against influenza has coincided so far with when the COVID-19 vaccines became available, improved and targeted messaging around co-administration of vaccinations against co-circulating viruses has the potential to improve coverage for COVID-19 for adults. Co-administration has economic benefits, can reduce attrition between preventative services, and can reduce structural and logistical barriers that impact adult vaccination^24,38,39^. While co-administration has the potential to increase COVID-19 vaccination uptake, other strategies such as provider messaging encouraging co-administration, follow-up with or scheduling the next vaccination appointment before the patient leaves may also reduce MoV^29^. Recommendations for co-administration of both COVID-19 and influenza vaccines need to consider eligibility, particularly for the COVID-19 vaccine which has exhibited non-seasonal patterns of transmission occurring in late summer^40^. Importantly, and while we demonstrated that targeting influenza vaccine recipients to also receive the COVID-19 vaccine can improve COVID-19 vaccination coverage to at least influenza vaccination levels, concurrent strategies should still be employed to achieve optimal vaccination coverage for both pathogens.

Our study had a few key limitations. First, the exact date of influenza vaccination was not captured in the survey; therefore, some adults may have received their influenza vaccine before the 2023-2024 COVID-19 vaccine was available, making co-administration impossible. While this is likely, as of September 2023 when CDC began monitoring influenza coverage for the season, only 9% of adults had received their influenza vaccine, with uptake increasing after that time^41^. Second, we applied a strict eligibility criterion where we classified participants who had a SARS-CoV-2 infection between September 2023 and January 2024 as non-eligible for a COVID-19 vaccine leading up to our survey in January. Since we did not collect the date of when they received their influenza vaccine, participants may have been eligible during this period if they received their influenza vaccine after September 2023 but before their SARS-CoV-2 infection. This may underestimate the number of participants eligible during the study period. Furthermore, vaccine contraindications, and receipt of the previously approved bivalent mRNA COVID-19 vaccine 3-8 weeks prior to the day of influenza vaccination may render adults ineligible for the 2023-2024 COVID-19 vaccine^42^. Strengths of the study include a population-representative assessment of vaccination uptake and co-administration among adults across key clinical characteristics such as prior SARS-CoV-2 infection, pre-existing chronic conditions, and access to healthcare. By collecting the date of prior SARS-CoV-2 infection, our study examined eligibility which allowed for a more conservative estimate of the impact of co-administration on improving COVID-19 coverage among adults.

## Conclusion

While both were low, seasonal influenza vaccination coverage was higher than that of the COVID-19 vaccination coverage during the 2023-2024 respiratory virus season. We observed higher uptake of the seasonal influenza vaccine among COVID-19 vaccine recipients during the same visit than the COVID-19 vaccine among influenza vaccine recipients, suggesting that adults who receive influenza vaccines are a potential target group to improve COVID-19 vaccination coverage through co-administration. Receipt of only the influenza vaccine among COVID-19 vaccine-eligible and willing adults represents an important missed opportunity for the COVID-19 vaccination. Concomitantly administered influenza and COVID-19 vaccines can be one strategy to increase vaccination coverage for both pathogens.

## Acknowledgements

The authors wish to acknowledge the survey participants and Ipsos for completing survey sampling and data collection.

## Ethical Approval

The study protocol was approved by the Institutional Review Board at the City University of New York (CUNY IRB 2022–0407). The Institutional Review Board determined that the study met the criteria for Human Subject Research Exemption based on collection of de-identified survey data only. Consent to receive survey invitations from the Ipsos KnowledgePanel is obtained during the recruitment process. Before responding to a survey, individuals must opt in and are reminded that their responses will be used for research purposes only and that they can choose not to answer any question.

## Data Availability

The datasets generated during and/or analyzed during the current study are available from the corresponding author on reasonable request.

## References

1. CDC. About estimated. Flu Burden. October 7, 2024. Accessed October 29, 2024. https://www.cdc.gov/flu-burden/php/about/index.html?CDC_AAref_Val=https://www.cdc.gov/flu/about/burden/index.html

2. CDC. Estimated COVID-19 burden. Centers for Disease Control and Prevention. August 12, 2022. Accessed October 29, 2024. https://archive.cdc.gov/www_cdc_gov/coronavirus/2019-ncov/cases-updates/burden.html

3. Anderer S. CDC: Overall deaths, especially from COVID-19, lower in 2023. JAMA. 2024;332(13):1043. doi:10.1001/jama.2024.17606

4. CDC. Preliminary estimated flu disease burden 2022–2023 flu season. Flu Burden. October 23, 2024. Accessed December 10, 2024. https://www.cdc.gov/flu-burden/php/data-vis/2022-2023.html

5. Jackson ML, Phillips CH, Wellwood S, et al. Burden of medically attended influenza infection and cases averted by vaccination - United States, 2016/17 through 2018/19 influenza seasons. Vaccine. 2022;40(52):7703–7708. doi:10.1016/j.vaccine.2022.11.011

6. Grohskopf LA, Ferdinands JM, Blanton LH, Broder KR, Loehr J. Prevention and control of seasonal influenza with vaccines: Recommendations of the Advisory Committee on Immunization Practices - United States, 2024-25 influenza season. MMWR Recomm Rep. 2024;73(5):1–25. doi:10.15585/mmwr.rr7305a1

7. DeCuir J, Payne AB, Self WH, et al. Interim effectiveness of updated 2023-2024 (monovalent XBB.1.5) COVID-19 vaccines against COVID-19-associated emergency department and urgent care encounters and hospitalization among immunocompetent adults aged ≥18 years - VISION and IVY networks, September 2023-January 2024. MMWR Morb Mortal Wkly Rep. 2024;73(8):180–188. doi:10.15585/mmwr.mm7308a5

8. Black CL, O’Halloran A, Hung MC, et al. Vital signs: Influenza hospitalizations and vaccination coverage by race and ethnicity-United States, 2009-10 through 2021-22 influenza seasons. MMWR Morb Mortal Wkly Rep. 2022;71(43):1366–1373. doi:10.15585/mmwr.mm7143e1

9. Lewis NM, Zhu Y, Peltan ID, et al. Vaccine effectiveness against influenza A-associated hospitalization, organ failure, and death: United States, 2022-2023. Clin Infect Dis. 2024;78(4):1056–1064. doi:10.1093/cid/ciad677

10. CDC. ACIP recommendations: Influenza (flu) vaccine. ACIP Vaccine Recommendations and Guidelines. October 22, 2024. Accessed October 31, 2024. https://www.cdc.gov/acip-recs/hcp/vaccine-specific/flu.html

11. CDC. ACIP recommendations: COVID-19 vaccine. ACIP Vaccine Recommendations and Guidelines. September 23, 2024. Accessed October 31, 2024. https://www.cdc.gov/acip-recs/hcp/vaccine-specific/covid-19.html

12. Kojima N, Taylor CA, Tenforde MW, et al. Clinical outcomes of US adults hospitalized for COVID-19 and influenza in the respiratory virus hospitalization surveillance network, October 2021-September 2022. Open Forum Infect Dis. 2024;11(1):ofad702. doi:10.1093/ofid/ofad702

13. Oordt-Speets A, Spinardi J, Mendoza C, et al. Effectiveness of COVID-19 vaccination on transmission: A systematic review. COVID. 2023;3(10):1516–1527. doi:10.3390/covid3100103

14. CDC. Flu vaccination coverage, United States, 2022–2023 influenza season. FluVaxView. September 6, 2024. Accessed October 28, 2024. https://www.cdc.gov/fluvaxview/coverage-by-season/2022-2023.html?CDC_AAref_Val=https://www.cdc.gov/flu/fluvaxview/coverage-2223estimates.htm

15. Lu PJ, Zhou T, Santibanez TA, et al. COVID-19 bivalent booster vaccination coverage and intent to receive booster vaccination among adolescents and adults - United States, November-December 2022. MMWR Morb Mortal Wkly Rep. 2023;72(7):190–198. doi:10.15585/mmwr.mm7207a5

16. Black CL, Kriss JL, Razzaghi H, et al. Influenza, updated COVID-19, and respiratory syncytial virus vaccination coverage among adults - United States, fall 2023. MMWR Morb Mortal Wkly Rep. 2023;72(51):1377–1382. doi:10.15585/mmwr.mm7251a4

17. CDC. Getting a flu vaccine and other recommended vaccines at the same time. Influenza (Flu). September 30, 2024. Accessed October 29, 2024. https://www.cdc.gov/flu/vaccines/coadministration.html

18. Pattinson D, Jester P, Gu C, et al. Ipsilateral and contralateral coadministration of influenza and COVID-19 vaccines produce similar antibody responses. EBioMedicine. 2024;103(105103):105103. doi:10.1016/j.ebiom.2024.105103

19. McGrath LJ, Malhotra D, Miles AC, et al. Estimated effectiveness of coadministration of the BNT162b2 BA.4/5 COVID-19 vaccine with influenza vaccine. JAMA Netw Open. 2023;6(11):e2342151. doi:10.1001/jamanetworkopen.2023.42151

20. Wagenhäuser I, Reusch J, Gabel A, et al. Immunogenicity and safety of coadministration of COVID-19 and influenza vaccination. Eur Respir J. 2023;61(1):2201390. doi:10.1183/13993003.01390-2022

21. Tzenios N, Tazanios ME, Chahine M. Combining influenza and COVID-19 booster vaccination strategy to improve vaccination uptake necessary for managing the health pandemic: A systematic review and meta-analysis. Vaccines (Basel). 2022;11(1):16. doi:10.3390/vaccines11010016

22. ACIP timing and Spacing guidelines for Immunization. July 22, 2024. Accessed October 31, 2024. https://www.cdc.gov/vaccines/hcp/acip-recs/general-recs/timing.html

23. Parker CE, Hause AM, Marquez P, Zhang B, Myers TR, Shay DK. Trends in the administration of COVID-19 vaccines with other vaccines in the United States reported to V-safe during December 14, 2020-May 19, 2023. Hum Vaccin Immunother. 2024;20(1):2361946. doi:10.1080/21645515.2024.2361946

24. Harris DA, Chachlani P, Hayes KN, et al. COVID-19 and influenza vaccine coadministration among older U.s. adults. Am J Prev Med. 2024;67(1):67–78. doi:10.1016/j.amepre.2024.02.013

25. Nguyen KH, Coy KC, Black CL, Scanlon P, Singleton JA. Comparison of adult hesitancy towards COVID-19 vaccines and vaccines in general in the USA. Vaccine. 2024;42(3):645–652. doi:10.1016/j.vaccine.2023.12.042

26. Be Sure with KnowledgePanel®. Accessed November 13, 2024. https://www.ipsos.com/sites/default/files/18-11-53_Overview_v3.pdf

27. COVID-19 vaccination provider requirements and support. October 17, 2024. Accessed December 10, 2024. https://www.cdc.gov/vaccines/covid-19/vaccination-provider-support.html

28. The World Health Organization (WHO). Methodology for the assessment of missed opportunities for vaccination. November 10, 2017. Accessed November 13, 2024. https://www.who.int/publications/i/item/9789241512954

29. CDC. Chapter 3: Immunization strategies for healthcare practices and providers. Epidemiology and Prevention of Vaccine-Preventable Diseases. July 10, 2024. Accessed November 19, 2024. https://www.cdc.gov/pinkbook/hcp/table-of-contents/chapter-3-immunization-strategies.html

30. U.S. Census Bureau. Explore census data. Accessed May 12, 2023. https://data.census.gov/table

31. Increase the proportion of people who get the flu vaccine every year — Data - Healthy People 2030. Accessed October 29, 2024. https://odphp.health.gov/healthypeople/objectives-and-data/browse-objectives/vaccination/increase-proportion-people-who-get-flu-vaccine-every-year-iid-09/data

32. CDC. Influenza vaccination coverage and intent for vaccination, adults 18 years and older, United States. FluVaxView. October 22, 2024. Accessed October 23, 2024. https://www.cdc.gov/fluvaxview/dashboard/adult-coverage.html

33. CDC. COVID-19 vaccination coverage and intent for vaccination, adults 18 years and older, United States. COVIDVaxView. December 16, 2024. Accessed December 20, 2024. https://www.cdc.gov/covidvaxview/weekly-dashboard/adult-vaccination-coverage.html

34. Weir JP, Gruber MF. An overview of the regulation of influenza vaccines in the United States. Influenza Other Respi Viruses. 2016;10(5):354–360. doi:10.1111/irv.12383

35. Salisbury D, Lazarus JV, Waite N, et al. COVID-19 vaccine preferences in general populations in Canada, Germany, the United Kingdom, and the United States: Discrete choice experiment. JMIR Public Health Surveill. 2024;10:e57242. doi:10.2196/57242

36. Rome BN, Feldman WB, Fischer MA, Desai RJ, Avorn J. Influenza vaccine uptake in the year after concurrent vs separate influenza and zoster immunization. JAMA Netw Open. 2021;4(11):e2135362. doi:10.1001/jamanetworkopen.2021.35362

37. Centers for Disease Control and Prevention (CDC). RESULTS FROM OMNIBUS SURVEYS ON VACCINATION RECEIPT, INTENT, AND KNOWLEDGE, ATTITUDES, BELIEFS, AND BEHAVIORS. October 2023. Accessed March 13, 2025. https://stacks.cdc.gov/view/cdc/148229

38. Equils O, Kellogg C, Baden L, Berger W, Connolly S. Logistical and structural challenges are the major obstacles for family medicine physicians’ ability to administer adult vaccines. Hum Vaccin Immunother. 2019;15(3):637–642. doi:10.1080/21645515.2018.1543524

39. Mehta D, Sun T, Wang J, Situ A, Park Y. Comparison of healthcare resource use and cost between influenza and COVID-19 vaccine coadministration and influenza vaccination only. J Med Econ. 2024;27(1):1190–1196. doi:10.1080/13696998.2024.2400852

40. CDC. COVID-19 can surge throughout the year. National Center for Immunization and Respiratory Diseases. September 26, 2024. Accessed December 22, 2024. https://www.cdc.gov/ncird/whats-new/covid-19-can-surge-throughout-the-year.html

41. CDC. Influenza vaccination coverage for persons 6 months and older. FluVaxView. October 23, 2024. Accessed December 20, 2024. https://www.cdc.gov/fluvaxview/interactive/general-population-coverage.html

42. Clinical guidance for COVID-19 vaccination. October 31, 2024. Accessed December 20, 2024. https://www.cdc.gov/vaccines/covid-19/clinical-considerations/interim-considerations-us.html

